# Ethnic inequalities during clinical placement: a qualitative study of student nurses’ experiences within the London National Health Service

**DOI:** 10.1101/2023.02.13.23285608

**Authors:** Chenel R Walker, Cerisse Gunasinghe, Hannah Harwood, Annahita Ehsan, Farah Ahmed, Sarah Dorrington, Juliana Onwumere, Paula Meriez, Nathan Stanley, Nkasi Stoll, Charlotte Woodhead, Stephani L Hatch, Rebecca Rhead

## Abstract

**Aim:** To understand how student nurse experiences on clinical placement, within NHS hospitals, differ for ethnic minority and White British groups.

**Design:** A qualitative thematic analysis with an inductive approach.

**Methods:** Data from semi-structured interviews with 21 London (United Kingdom) hospital-based student nurses were examined using thematic analysis. Participants were interviewed as part of the *Tackling Inequalities and Discrimination Experiences in health Services* (TIDES) study and asked about their experiences during clinical placement.

**Results:** Five main themes were identified: 1) Role of mentors, 2) Discrimination and unfair treatment, 3) Speaking up/out, 4) Career progression, and 5) Consequences of adverse experiences. All themes were linked, with the social dynamics and workplace environment (referred to as ‘ward culture’) providing a context that normalises mistreatment experienced by nursing students. Students from ethnic minority backgrounds reported racism as well as cultural and/or religious microaggressions. While being valued for their race and ethnicity, White British students also experienced discrimination and inequity due to their age, sex, gender, and sexual orientation. Students from both White British and ethnic minority groups acknowledged that being treated badly was a barrier to career progression. Ethnic minority students also noted that the lack of diverse representation within senior nursing positions discouraged career progression within the UK National Health Service (NHS).

**Conclusion:** These initial experiences of inequality and discrimination are liable to shape a student’s perspective of their profession and ability to progress within nursing. The NHS is responsible for ensuring that student nurses’ developmental opportunities are equal, irrespective of ethnicity.

**Impact:** Ward culture is perpetuated through the action of others to normalise mistreatment and concurrently disadvantage ethnic minority students making them feel unvalued, which in turn impacts both staff retention and career progression within the NHS. Training assessors should be aware of the existing culture of discrimination within clinical placements and work to eradicate it.

## 1. Introduction

Career progression for ethnically minoritised staff in the National Health Service (NHS) is often hindered (1). They have less chance of being shortlisted for jobs and being invited to non-mandatory training, yet have a higher chance of being penalised at work than their White British colleagues (2,3). As a result, despite comprising over 20% of the NHS workforce in England (45% in London Trusts (5)) (3,6), only 8% of NHS senior leaders are from an ethnic minority background (3). Ethnic minority staff are also more likely to experience bullying and discrimination from both colleagues and managers (particularly in London Trusts) (7). Such incidents are associated with poor mental and physical health outcomes, which in turn can lead to increased sick leave and negatively impact overall quality of life (7,8). These discriminatory experiences compound the already highly stressful work environment that nursing students are exposed to during their training (11,12), ultimately resulting in high staff attrition among student nurses (11,12).

## 2. Background

For student nurses, clinical placements are a necessary requirement of their training and crucial to their professional development. During clinical placements, student nurses gain crucial first-hand experiences in providing patient care and working alongside others within health and social care organisations. It offers them the chance to build confidence, gain practical experience and work compassionately within a multidisciplinary team (4). For ethnically and racially minoritized nurses, experiencing discrimination during these placements could impact future career progression by dispelling confidence and discouraging attainment of senior roles (13). For ethnically minoritised student nurses, it may also be their first experience of systemic disadvantage as an employee/staff member within healthcare services (14,15).

One theory that may explain such occurrences is the conceptual framework of marginalisation specifically relating to nursing education and practice (16,17). It has been proposed that there are seven fundamental aspects of marginalization: (1) intermediacy, (2) differentiation, (3) power, (4) secrecy, (5) reflectiveness, (6) voice, and (7) liminality, that either foster or impede diversity in the nursing workforce (16,18,19) as well as in the health care of diverse communities (16). Further, intersectionality theory may also offer insight into what systems of inequity student nurses may be exposed to during their education and training. Intersectionality theory offers a framework for understanding how multiple personal, political and social identities often being negotiated at any given time, intersects with discrimination and systems of oppression (8,20).

For nurses, the social dynamics and workplace environment they must navigate is often referred to as ‘ward culture’ (21). Not being able to adapt adequately and/or be socially accepted on a new ward during placement can be highly detrimental, potentially resulting in harmful workplace experiences and/or stunted professional development (22). Given how formative clinical placements are for student nurses, it is particularly important that they be allowed to integrate and adapt to their new care environment (23), benefit from the socialisation that occurs throughout the course (24) and avoid experiences associated with *maladaptive* ward culture. An example of how maladaptive ward culture can manifest is when mentors (who provide support for student nurses throughout their placements) exploit the mentor-mentee relationship. Mentors have the power to foster non-supportive relationships with students, knowingly or unknowingly enforcing hierarchical workplace structures which contribute to a toxic learning environment (25). This could be evidenced by mentors providing menial tasks outside the remit of the student nurse’s current role or withholding beneficial opportunities.

While all students are exposed to ward culture, there may be an additional hidden curriculum (i.e., unspoken or implicit values, behaviours, procedures, and norms that exist in the educational setting (26)) - one that only ethnically minoritised nurses are excluded from during training. The hidden curriculum has the potential to impact how student nurses are socialised into their workplace, and if ethnically minoritised students are not privy to this then it can impact their development (27). Students who do not face this dual hidden curriculum (e.g., White British students) may be unaware that it even exists, as student nursing research commonly overlooks this (28), and accidentally feed into it as their careers progress at the expense of their ethnically minoritised colleagues. The existence of a hidden curriculum within nursing education specifically goes against the NHS constitutional values of *“Everyone counts”* and *“Compassion”* (29). These values are required to be modelled by mentors, as they are integral to the NHS (30). However, these professionalisms and values may not be extended to all students.

## 3. The study

### 3.1. Aims

This study aimed to understand how clinical placement experiences diverge for students from ethnic minority (from both British and migrant backgrounds) and White British groups. The two objectives were (i) to understand the role of mentor support, and (ii) to explore inequities related to career progression.

### 3.2. Design

This study presents a sub-analyses of qualitative interview data from the mixed methods Tackling Inequalities and Discrimination Experiences in Health Services (TIDES) study. The TIDES study aims to understand how discrimination experiences contributes to inequalities in health and health services. Full details of the methods and sample description were previously reported (7,8).

### 3.3. Participants and sampling

Participants were recruited as part of phase one of the TIDES study (7). Participants of this study initially completed a survey designed to assess their experiences of working in the NHS and the impact such experiences had on their health and wellbeing (n = 931, detailed in Rhead et al. (2021)) (7). Survey participants who gave consent to be re-contacted were also purposefully sampled for interviews (31). A total of 225 survey participants (including student nurses, midwives, and healthcare assistants from diverse racial and ethnic groups) were contacted by email (up to three emails were sent per person) and invited to take part in an interview. Those who expressed interest were sent a participant information sheet which included details on confidentiality and how to withdraw from the study.

From the 225 staff contacted to participate, 48 staff took part (6 health care assistants, 21 student nurses, 10 entry level nurses and 11 mid or senior level nurses). The 179 people contacted who did not take part either did not respond or did not have the time to participate. On completion, participants were offered a £15 voucher as a thank you for their participation.

Inclusion criteria for the current sub-study analysis were: 1) Student nurse and 2) Previous experience of a clinical placement. Those with no experience of clinical placement, or those employed as a healthcare assistant or qualified nurse were excluded from this sub-study analysis. Once these criteria were applied, of the 48 interviews conducted, 21 were eligible for this study. Of these 21 eligible participants, four were male, 10 were from an ethnic minority group and four were migrants (all migrant participants were from an ethnic minority group). Figure 1 outlines the selection process.

**Figure 1:**
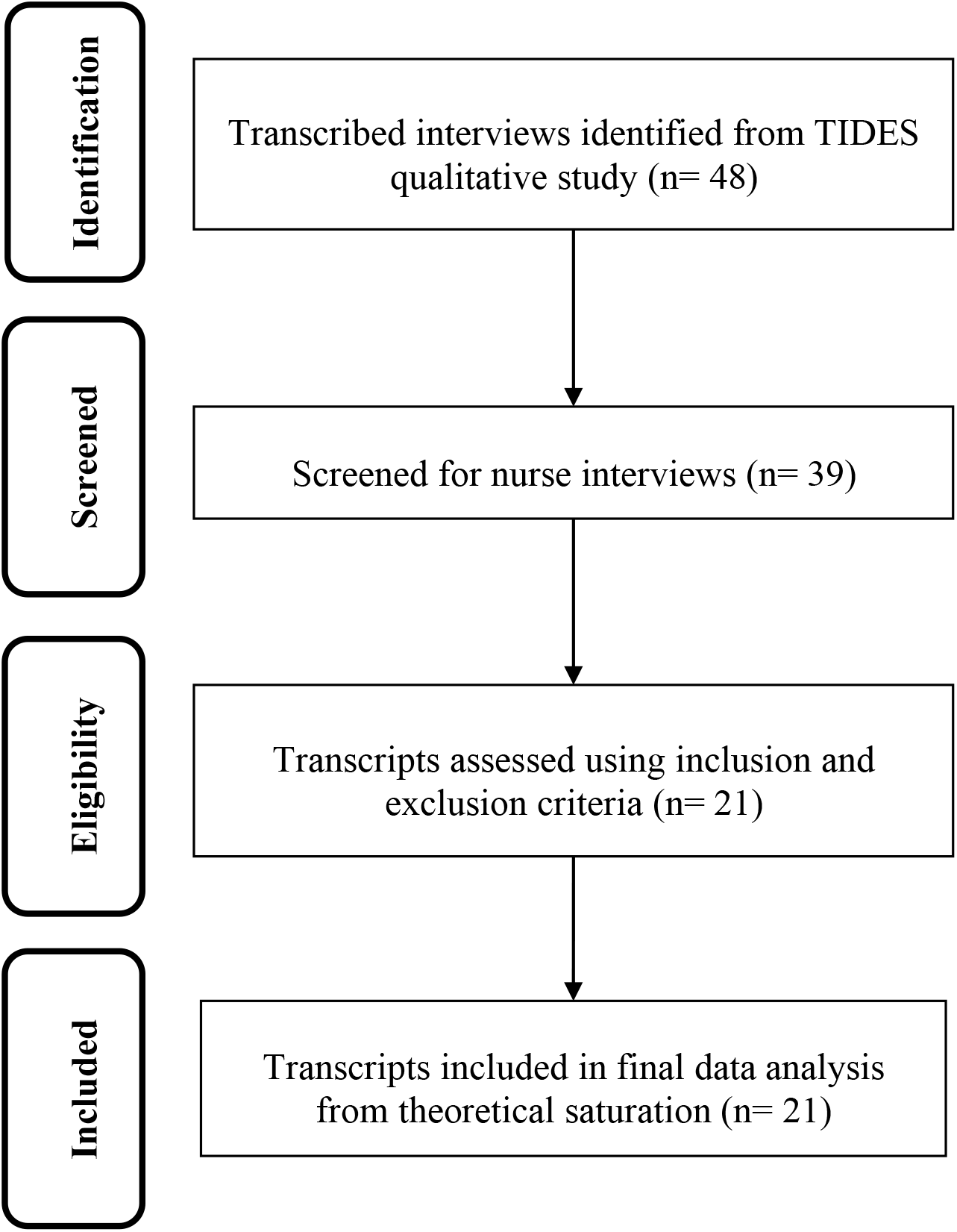
Transcript selection process

**Figure 2:**
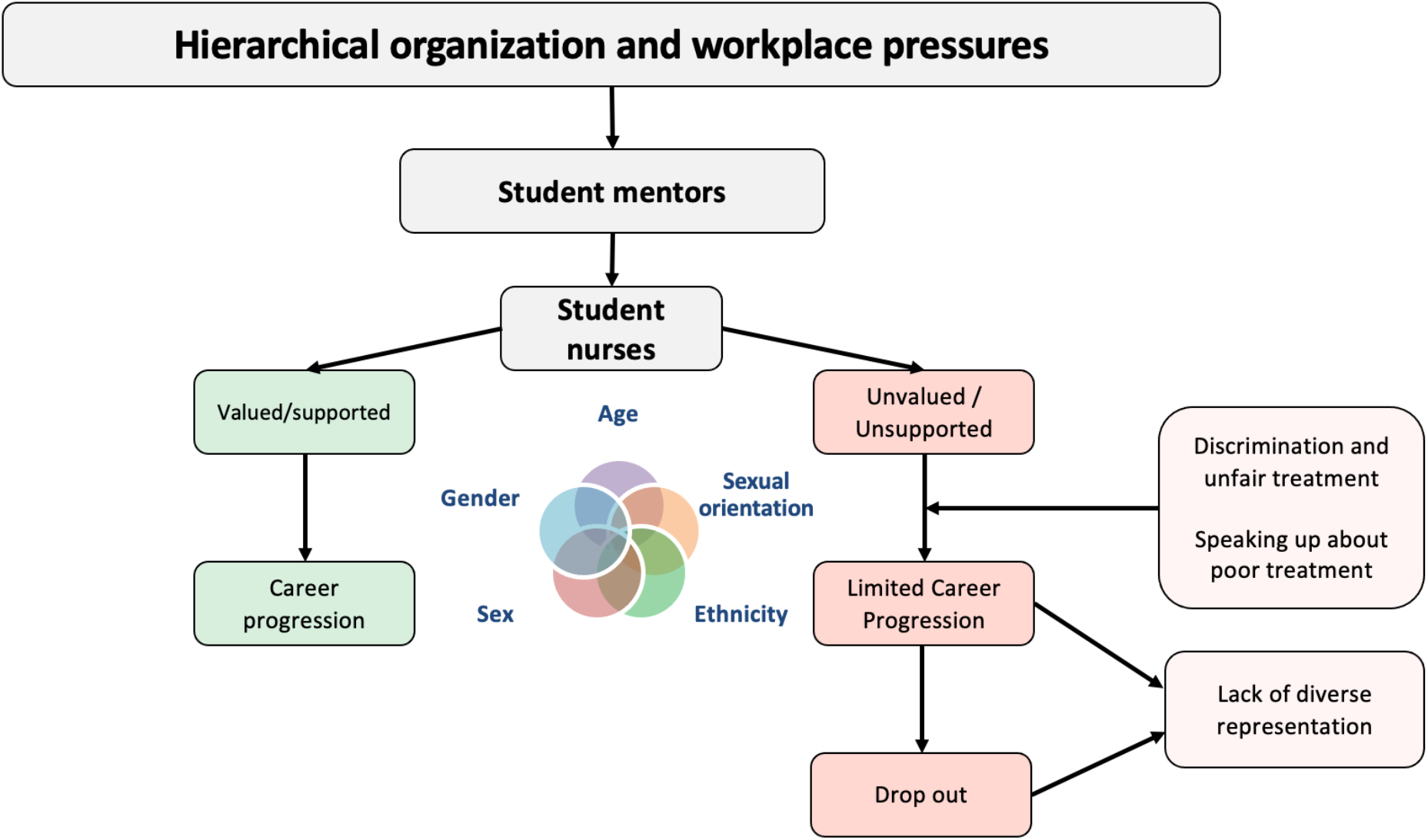
Overview of ward culture, feeling valued and supported, and career progression

### 3.4. Data collection

HH informed participants that the interview topics would explore work environment, witnessing, experiencing, and reporting bullying, harassment, or discrimination as well experiences of training and support and explained her role within the research. Interviews were conducted between January 2019 and February 2020. Participants were assigned a unique ID number and provided informed consent prior to interview (5).

Data were collected via individual semi-structured in-person or telephone interviews (based on participant preference), completed by HH which lasted up to 60 minutes. Each participant had one interview which was audio-recorded. HH ensured she was in a room alone and asked the participants prior to conducting the interview whether they were alone and able to begin. Interviews were transcribed verbatim, using an external transcription service.

#### Ethical considerations

Ethical approval was granted from the university’s Research Ethics Committee: HR-17/18-462, as well as NHS ethical approval Project ID: 230692.

Prior to interviewing participants, written informed consent was obtained. Participants were provided with information sheets, outlining the study purpose, data handling, the topics being covered, their right to discontinue the interview and the time frame up to which they could withdraw their data. Participants were able to withdraw their data from the study up until 1 month after interview, however, no participants requested to withdraw their data (5). Participants gave consent for their interview to be audio recorded. Quotes are labelled with participant gender and ethnicity (ascertained from the TIDES survey responses).

To protect anonymity, identifying information were removed or redacted from the transcripts. Participant demographics and transcripts were password protected and saved securely on a digital platform with restricted access to key researchers. Lastly, no identifiable quotes have been included within the report to protect participant anonymity.

### 3.5. Data analysis

Data were analysed using thematic analysis (32) supported by *NVivo 20* software (33). This approach follows a reflexive paradigm rather than prioritizing data saturation (34). This approach was most appropriate as the sample of student nurses was limited in the larger study, and we were interested in emerging themes that centred around participants’ lived experiences. Primary data analysis was conducted by CRW (who analysed 12 transcripts), their code was reviewed by co-authors. The remaining nine transcripts were coded by co-authors CW, NS, CG, FA and JO. These codes were compiled and reviewed by CG.

### 3.6. Rigour

Throughout this study the researchers respected *Lincoln & Guba’s* criteria for assessing rigour (35). Coding was reviewed by co-authors to enhance dependability and trustworthiness of the analysis. PM, a peer researcher working on the TIDES project with a background in nursing, reviewed and contributed to the paper. Throughout the study co-authors met to discuss the themes identified, this enriched the study’s credibility as the researchers were confident with how the interpretations were presented.

### 3.7. Reflexivity

Interviews were conducted by a TIDES research assistant, HH. HH identifies as a young White British female with previous experience gathering qualitative data. Twelve transcripts were coded and analysed by the master’s student and first author (CRW), who identifies as a young, Black, woman. CRW has previous experience with research for her undergraduate degree in Psychology and more recently as a postdoctoral student.

To consider reflexivity, CRW has previously witnessed and experienced racial micro-aggressions – these are defined as small events that are often ephemeral and hard to prove and events that are covert, often unintentional, and frequently unrecognized by the perpetrator that occur wherever people are perceived to be “different” (36). Witnessing discrimination first-hand within the NHS towards ethnic minority staff may have influenced how she interpreted the data. CRW holds the belief that micro-aggressions could be an issue affecting pre-qualified nurses, and there is a possibility that this could have impacted how she attached meaning to the excerpts. Nevertheless, the analysis of the data and the review of this paper were conducted by a research team with a diverse racial and ethnic composition.

## 4. Findings

As shown in Table 1, Five main themes and subthemes related to the study aims were identified from participant transcripts: (1) role of mentors, (2) discrimination and unfair treatment, (3) speaking up/out, (4) career progression and (5) consequences of adverse experiences.

**Table 1:** Themes and subthemes.

| Themes | Sub-themes |
| --- | --- |
| *Role of mentors* | *Feeling valued and supported* |
|  | *Lack of value and feeling unsupported* |
| *Discrimination and unfair treatment* | *Intersectionality and inequalities* |
|  | *Abuse, bullying and harassment* |
| *Speaking up/out* |  |
| *Career progression* | *Diverse representation* |
| *Consequences of adverse experiences* | *Creating distress and dissatisfaction within working relationships and environments.* |
|  | *Impact on patient care* |

### 4.1. Role of mentors

Participants spoke of varied experiences of mentorship, professional skills development and acquiring knowledge while on clinical placements. This was attributed to contextual/organisational factors, as well as the individual mentor’s interpersonal skills. Subthemes within this theme were labelled as *Feeling valued and supported* and *Lack of value and feeling unsupported*. The consequences of adverse experiences with mentors and within the organisational structure and culture are presented in theme five.

#### Feeling valued and supported

Some participants, who were mostly White British, reported positive experiences and frequently disclosed they felt valued by their mentor and the wider team across clinical placements.

…*I think em, every mentor I’ve had, I’ve got on with*… *Em, quite well*… *And they’ve, they’ve always been really, really, helpful and*… *Supportive. So, I’ve never, never had a problem with them. (Female, White British)*

*I think it depends on, it depends, it definitely depends on where you are. So, for example, I, I felt like, when I was on my community placement, I, I really enjoyed it, because I was always working alongside the nurse. And we were, like, always doing things together. Helping each other, um*… *(Female, Black Other)*

Some White British students had positive experiences while on clinical placements because they were valued by patients for their whiteness:

*Um, also I think I’m White and I’m British and, um, with certain patients that seems to…that seems to seem like a good thing. Um, older patients seem to like the fact that I’m British and I’ve heard them, yeah, moaning about um patients from other…nurses from other countries before. (Male, White British)*

*I think because I’m White British sometimes patients think it’s okay to talk to me about, like I remember one patient saying to me, she was being quite racist about some of the other staff members and I think she just assumed that because I was White and from Britain that I felt the same as her, which I didn’t, so I felt really uncomfortable but I kind of said to her, ‘We don’t kind of talk about that here, like it’s not okay to talk about people like that’, but it was really like uncomfortable for me and I felt I couldn’t really, I didn’t want to tell anybody about it but yeah, I felt quite uncomfortable going back in and looking after that patient again. (Female, White British)*

This suggests that, for student nurses, feeling valued by their mentors is essential for a successful placement with positive professional outcomes.

#### Lack of value and feeling unsupported

Conversely and more prominently to the above, most of the participants (regardless of ethnicity) experienced an absence of mentorship, felt unvalued and unsupported (e.g., allocation of menial tasks, being used as labour, perceived as inexperienced, due to their “student status”). This was also witnessed towards other nursing students, lower band staff (healthcare assistants) and staff groups (bank and agency staff). ‘Hierarchical organisation and workplace pressures’ as identified by Woodhead et (2022) whereby workforce hierarchies create and maintain inequalities experienced by trainees and junior staff, were mechanisms and experiences of organisational culture that also resonated for this sub-sample of student nurse participants:

*I felt like they didn’t really wanna involve us in um, key tasks that was fundamental to our learning, for example um medication. Um they are really sceptical about our knowledge in that area because we’re all quite new into the training. (Female, Asian)*

*And when I say power it’s just we…we see a lot and we work with a lot of different nurses on the ward, or healthcare workers, so you get to know, like, little bits from a lot of different people. So you, kind of, possess, like, a lot of information that, you know, maybe other people wouldn’t necessarily hear or know, erm, yeah, I just…yeah [short pause], I just…I just think that, erm, yeah, we’re probably just seen as a bit untrustworthy. (Female, White British)*

*But then I did have one, actually, in my last one placement who, she wasn’t very helpful, she didn’t really explain anything to me or delegate to me. And I didn’t really know where I stood, like, what I was meant to be doing, so I didn’t have to talk to her. (Male, Black African)*

As can be seen here and identified within other participant transcripts, White British and ethnically minoritised students had similar explanations that organisational hierarchy and culture appeared to be continuously reinforced by their student mentors and senior colleagues. As seen in the last quote above and the quote below, work pressures and interpersonal style of mentors also contributed to negative training experiences for participants while on clinical placements:

*“I think sometimes maybe they just don’t really want to be a mentor or a teacher, maybe it’s something they’ve kind of been lumbered with and I can see it’s like another, everyone’s very busy, I can see it’s like maybe quite difficult for people to have an extra student but I think if they’ve got a good student they’re with the student is a big help to them as well. I think just maybe not explaining things properly or maybe not really answering questions, things like that, maybe just, I know everyone kind of has a different learning style but I like to feel kind of comfortable with that person to be able to ask questions and things and not feel judged about asking questions but I think some mentors can make you feel a bit stupid if you ask questions and things like that*…*” (Female, White British)*

Many ethnically minoritised students did not explicitly report feeling valued or supported. Instead, they recalled experiences of unequal treatment. Examples included mentors excluding them from learning opportunities, and occasions where mentors raised their voice to their student. In addition, some participants spoke of how minoritised groups were perceived differently by mentors, which led to discriminatory attitudes and treatment:

*Um, it seems to be mainly Black nurses, um. Either West Indian or African nurses. They seem to say that they’re…the attitude is that they’re not as good and also that they are, um, superior and perhaps bolshie… Yeah, um. Someone I was working with, um before said, um, she said, she was working on a ward with a lot of, um Black African nurses and she said um, she said that she didn’t like working with them and I asked why, and she said that they had a lot of attitude and then she used the phrase “Black attitudes” and she said similar sort of things to what the patients would say. (Female, Asian)*

This also illustrates how ethnically minoritised student nurse participants’ experiences of feeling unsupported and not valued by mentors overlaps with the second main theme of discrimination, in particular exposure to racism or microaggressions, and unfair treatment which is presented in more detail below.

### 4.2. Discrimination and unfair treatment

Within this theme, student nurse participants spoke of discriminatory attitudes and unfair treatment relating to personal and social identities. Abuse including exclusionary behaviours, bullying and harassment were also experienced and witnessed by student nurse participants. These were perceived as upholding organisational hierarchy and/or socio-cultural norms in addition to an exposure to an individual’s own prejudices and stereotypes.

For some participants, exposure to discrimination and unfair treatment was interlinked with their lower position in the organisational hierarchy and the sub-theme of lack of value and feeling unsupported:

*I feel like sometimes I’m a student nurse, it depends on, um, different, so, different, different types of people you’re working with. So, um, as a student nurse, I feel like we’re not*… *sometimes we’re not valued, um, enough. Um, and sometimes that people underestimate, like, what we can do in terms of, like, um, like, caring for our patients. Even though we’re still learning, we feel like, um, sometimes we’re discriminated against because we’re, like, we’re always told, like, there’s a student and we need to do like all the, like, things, like, for example, as a student nurse I feel like it’s important for people to be able to learn and experience things as a student nurse rather than just like a healthcare assistant. So sometimes you can find that they use you more as a healthcare assistant rather than actually being there to, like, learn from nurses. (Female, Black Other)*

#### Intersectionality and inequalities

Many participants spoke of discrimination and unfair treatment associated with protected characteristics such as age (being perceived as young), gender (inequalities experienced by male student nurses), sexual orientation and/or paternity/maternity:

*Yeah, urm, the thing that happened with me which I was [inaudible 00:13:46] because, urm, the way I looked. I looked [inaudible 00:13:51] petite and, urm, look young. They don’t really, urm, trust me to do, urm, certain things on the ward. (Female, Black Caribbean)*

*When I first went on placement, I picked up really quickly that most of the…most of my colleagues, like, felt quite hostile towards LGBT people, so I just decided at that point, like, just to not talk about…just to, like, keep my identity to myself while I was doing the course. (Female, White British)*

*Um, yeah I have actually. I have a friend, um, working in another ward, she’s actually trying to get pregnant as the moment, and she…it’s something that she’s very excited about. So she mentioned it to her ward sister, um, and the ward sister kind of said to her, I hope you are trying to get pregnant because then we won’t hire you*… *Um, and she said it as a half joke, um, but my friend was sure it wasn’t a joke, um, and she spoken to her since. I mean, they haven’t spoken to each other. So now she’s 100% sure it wasn’t a joke, um, and it is things like that that make, makes me very wary about what I say exactly and to whom I say it to, um, because I think there can definitely be that kind of discrimination around. (Female, White British)*

In addition to exposure to the discrimination as described above, participants from ethnic minorities reported discrimination connected to their race, religion, culture and/or language:

*Overall, attitude which could be cultural, or other people understanding their culture and how they do things, and how they say things. But so far, touch wood, I haven’t really experienced others … Yeah. I would say that generally the idea in the work environment is that, certain ethnic backgrounds are a bit lazy, and they don’t have the right attitude, but I find that more with the older, more qualified staff who may be put out, and some mature students. (Female, Black Caribbean)*

*Um, so, they’d go out after work with the White English, like, they’d include her in as well. Like, they’d go out for drinks whatever. But the black student nurse was never*… *wouldn’t be invited. (Female, Black Other)*

#### Abuse, bullying and harassment

Student nurse participants spoke of experienced bullying, harassment, and other mistreatment that they normalized as being part of nursing wards:

*So there’s a culture of bullying and harassment. I have experienced this first-hand in every way. Because there are harassment and bullying in different forms, apart from what was on paper. Especially where it, it is very difficult to prove. (Male, Black African)*

*Um. From fellow staff, I think because, I mean, you kind of get a bully, and someone’s physically it… abused quite a lot by the patient, and you kind of develop quite a thick skin towards that. But if it’s people within your own team then, you know, it definitely has more of an impact. (Male, White British)*

The neutral labelling of unfair treatment normalised some of these experiences and allowed it to continue, which reinforced the cycle of mistreatment. Further, this also was true for participants who spoke of abuse by mentors:

*Erm, I was actually speaking to one of my colleagues today and she was telling me a story of, in her last placement that her mentor was being sexually inappropriate, and she reported it to the University, erm, err, but nothing was really done about it because they basically said, “Well if you’re gonna take this any further that you need to…” you know, go to, n-not court but there needs to be, you know, like, this serious, erm, report, you know, and then an investigation, and then a hearing. And I think cause of where she’s currently at that wasn’t something she was prepared to do, so it hasn’t really gone any further. (Female, White British)*

*She’s trying to catch up with me, and she’s trying to make sure she’s fulfil her duties as a mentor. Everything’s she’s trying to do at the very last minute. And, the funny, the thing is I actually had an altercation with her on one of the night shifts. And. Something happened on the ward, um something happened. And we exchanged words and I told her, don’t ever speak to me like that. And she wasn’t really happy about me telling her that, I’m like, whatever you saying don’t shout at me, don’t try to raise your voice at me. You know, just because I look a certain way doesn’t mean I cannot actually, you know, speak back at you, so. (Female, Asian)*

### 4.3. Speaking up/out

This theme illustrates how adverse experiences with mentors and during clinical placements were managed by the student nurse participants including the dilemma or negotiation of tensions (personal and structural) that they were having to navigate when exposed to discrimination and unfair treatment. Few participants spoke of advocating for themselves or others in reporting adverse experiences like unacceptable and inadequate mentorship, bullying, discrimination, harassment and unfair treatment:

*So if you’re hearing them being disrespected, or if you’re seeing them being mistreated then you have, like, a moral responsibility to do whatever you can to try and stop that. Even if the systems then designed to avoid having to take what you’re saying on board, you have to just do whatever you can within the limits of what power you’ve got. (Female, White British)*

*Um, whether to report something. No. It depends. Sometimes, um, if it’s, if it’s, like, bullying or harassment and it’s*… *I, I would always report it. I don’t think anything would actually influence it. (Female, Black Other)*

One student mentioned not reporting poor experiences because of their student status:

*I’ve never spoken about it* … *because I think it’s just something you brush under the carpet and made me question how many times it happened to me in the past but because it’s been my profession, I haven’t really thought about it and it’s just more pronounced now that I’m a student. (Female, White British)*

Some participants also recalled that they felt more comfortable, safe and had more positive experiences of reporting adverse experiences to their academic institution than the NHS manager or Trust, while on clinical placements. For others, there appeared to be a negotiation between managing their own psychological safety; the safety of others (colleagues and patients); perception of, or actual repercussions for their career and not colluding with the organisational culture or being a bystander:

*I think I’d feel really unsure, I think I’d find it really difficult if it was say the manager that was discriminating, if I’d witnessed the manager discriminating against another member of staff, I think I’d find that really difficult because on one hand that person is your manager at the same time but I don’t think it’s right to not say anything and for that to not be reported but I think I’d be worried about if the manager like found out and I’d lose my job and things like that. So, I think I’d find that quite tricky. Maybe I’d find it a bit easier if it was like a patient or a staff member, actually no, I think I’d find it easier if it was a patient maybe being discriminatory towards a member of staff, I think I’d find that easier to kind of escalate whereas if it was any staff member maybe being abusive towards a patient or discriminatory I think that would be a bit more difficult. I think in that sense maybe safeguarding would be probably appropriate if it was a staff member maybe being discriminatory towards a patient. But yeah, I think that’s quite a hard one. (Female, White British)*

*I…I, again, I would definitely support them in doing that but I wouldn’t want to…it-it’s difficult as a student because you are on this tightrope of wanting to do right by everyone, but also having to think about yourself and the repercussions that it can have on, like, your experience*… *So, for example, coming back to the hostility with mentors, if you have a mentor who’s not very good and not very supportive, if you said something to them they…they could punish you in some way, erm, by, for example, not assigning skills, or not assigning your hours, or not doing your mid-point, or whatever it is they…they have the power. So I think you have, as a student, you have to be really careful. And I think that would…is probably what holds me back the most, erm, in, not…not reporting bad practice cause I think I would always do that, erm, but it is definitely something that, err, yeah, I find…I find really hard to do. (Female, White British)*

*I’d still report but I’d probably be utterly terrified about doing it, um, and with my anxiety I’d probably lose sleep over it. Yeah, it wouldn’t put me in a good space with it, um, or because I didn’t have a mentor then at that stage, um, I wasn’t able to talk about it with them, um*… *(Female, White British)*

Reporting discriminatory treatment appeared to be mishandled in some cases. For example, one ethnic minority student reported poor training on multiple occasions, but it did not result in change:

*So, with the training, yes, we get the training, and they will tell you channels for reporting. But I have put in, like, three or four grievances in the last several years. They’ve all been mismanaged even where there is overwhelming evidence beyond reasonable doubt. They have all been mismanaged and said, this is not found, that is not found (Male, Black African)*

### 4.4. Career progression

Career progression was another prominent theme, with discussions centred on the barriers to career progression within the NHS. Career progression was related to organisational culture: some ethnically minoritised nurses appeared to be more affected by ward culture and did not have positive mentors to help mitigate its negative effects.

Both White British and ethnically minoritised student nurses shared the belief that treating student nurses poorly (and witnessing qualified staff being treated unfairly) would result in increased student nurse dropout rates. Student nurses saw this as mentors creating barriers within their nursing placements and at an early stage of their career.

*Um, but yeah so for sure I think they can, and it’s such a shame because they’re so lovely people and they would be amazing nurses when they get to the end of their career, but they’re just getting treated like rubbish and that’s why they don’t want to stay there anymore. (Male, White British)*

*You know, I mean, and I know that if I had other people have had that experience, and I know for a fact if you’re doing a survey you need to look at the drop off rates that happen on the course, and once people start going on placement. (Female, Black Caribbean)*

One potential outcome of suboptimal mentorship experiences during clinical placements is the development of apprehension about future career progression. For example, one student reported that a having a *“bad”* previous experience on placement with mentors affected student confidence throughout their professional journey. Negative experiences during clinical placements may feed back into negative experiences that become normalized as part of organisational structure and culture. White British nursing students, who feel more supported and valued by their mentors, may advance in their careers and unknowingly reinforce ward culture in the future.

#### Diverse representation

Lack of diverse representation was problematic for some ethnically minoritised participants and their perceptions of career progression.

…*In my previous role, I would say that I have been held back from progressing, and I think it’s possibly because I’m from a BAME background*… *where I could have … that certain roles weren’t made possible. Once I felt like I was thrown under the bus when they were trying to investigate someone, but they needed a scapegoat, type of thing*…*Yeah. But mainly the fact that I possibly could have stepped up into roles many years prior to when I did, and they didn’t make it possible, or they made it more difficult for me to …That’s okay. Even the nursing degree. I could have done these two years prior, but it’s okay because I’m doing now and then I’m finished next month*…*Yeah. So, because I was in admin, I used to get that, “Oh well, it’s not related to your role”. (Female, Black Caribbean)*

*Um I mean it’s too early for me to tell at the moment, but I do get moments where I don’t really see um enough people like um, who are [two minoritized groups] at the same time doing what, you know, in the profession that I’m aspiring to get into. So I can feel like there could be some barriers stopping people progress within that field because of that. (Female, Black African)*

Ethnically minoritised student nurses recognised a lack of diverse representation in senior positions and highlighted that this is a potential limitation for progression with the nursing profession.

### 4.5. Consequences of adverse experiences

Where there were adverse experiences with mentors and because of wider organisational culture, participants spoke of how this impacted their own well-being and that of colleagues, as well as patient care.

#### Distress and dissatisfaction in the workplace

Some of the student nurse participants spoke of how exposure to unfair treatment impacts emotional well-being:

*Yeah. For example, I was changing a patient one time with, um, one of the healthcare assistants. And, um, she kind of, she kind of, she said to me, oh, um, have you ever changed a patient before? Like, she goes, and you haven’t*… *you’ve got younger brothers and sisters. I said to her, um, actually, I’m a mum. I’m a young mum, by the way. And she goes to me, um, she just said, she literally just said, she turned around and said, I can’t believe that you’ve got a kid. You can’t even*… *you don’t even know how to change a patient. You probably don’t even know how to change yourself. And then I just looked at her and I was like, oh, that’s not*… *like, that’s not really nice. I walked away. And then I just started*… *I couldn’t hold my tears back because, like, I know that she’s doing this because of my age. And then I know that she’s treating me like this*… *(Female, Black Other)*

*And I’ve witnessed um certain student colleagues just in tears because they’ve just, they’ve just been broken down by their mentor, you know. (Male, White British)*

*In one of the hospitals I was in, like, there was a rumour that a gay man, member of staff on another ward had, like, been bullied and had gone off sick. (Female, White British)*

For some participants, the organisational culture and work pressures created despondency and unhealthy team dynamics and working environments:

*I imagined like nurses would really care for each other and kind of be there to help each other out if they need it but actually, when it comes down to it, I think obviously time constraints and like pressures in the NHS don’t help but a lot of nurses I’ve encountered don’t often seem to be wanting to help each other out or can often kind of be a bit nasty to each other, which isn’t good*. (*Female, White British)*

*I would say that, people that complain, if anyone has bothered to make a complaint, be it bullying, harassment, they*… *it needs to be taken very serious. People are*… *because probably some people has misused this in the past. They don’t take complainants serious anymore. So, they go to some checkbox list. Have they harassed you on your kind of sexuality? No. On your race? No. On your this? No. Was it really their character then is not found. And such unhappy people carry on working in the NHS. And when people are unhappy, most people feel they are being, I would say it’s*… *Cheated is not the word. If people think they are being trampled upon, they will seek to trample on other things that are of value to those people. (Female, White British)*

*I didn’t want to stay there. I was like*… *You know when you just don’t want to accept it because you hear such good stuff about [HOSPITAL]? And you just think, maybe it’s me, you know. I’ll get better at my job. And you get better and you get better and then it’s like, no, it’s still the same. They’re still being like this. I’m leaving. (Female, Black Other)*

#### Impact on patient care

Some participants spoke of the consequences for patient care when staff experience lack of support and of no value to those within the organisational hierarchy:

*So, an-and I think as well that also impacts, erm, patient experience, you know, if nurses don’t feel valued or in…in any member of the team don’t feel valued, or don’t feel like they’re supported by management then I think that’s, you know, inevitably going to affect the way patient’s are cared for because of the, you know, mindset that staff are in. (Female, White British)*

While on clinical placements, some student nurse participants also witnessed colleagues’ discriminatory attitudes and unfair treatment towards patients, many of whom were from racial and ethnic minoritised backgrounds:

*Um, so like it really, really affected me seeing that. Like I even had one patient who, um, was kind of crying when I was about to leave my shift and she was saying please don’t leave, like please stay overnight because I’m scared that when you come here tomorrow morning I won’t be here. (Female, White British)*

*Is that from what I’ve, like*… *Okay. So, okay, so, like, the sickle cell patients, um, a lot of the time, around pain, especially. Like, we’ve had sickle cell kids on our ward. Um, and nurses have been like, oh, they’ll be, they’re just faking it. They’re fine. They can exaggerate. They know exactly what to do to get medication. So they have that perception rather than being like, no, actually, it’s a really bad pain. It’s worse than labour pain. That’s what it’s been described as. They need their medication right now*… *(Female, Asian)*

## 5. Discussion

This study conducted much needed research to understand how the experiences of student nurses on clinical placement, within NHS hospitals, differ for ethnic minority and White British groups. After analysing interview transcripts from 21 student nurses working in London Trusts, five key themes on how nursing students experienced their clinical placements were found: (1) the role of mentors, (2) discrimination and unfair treatment, (3) speaking up/out, (4) career progression and consequences of adverse experiences.

Overall, student experiences of the workplace and organisational culture were found to determine whether participants had positive or negative experiences with mentors. Student nurse participants from ethnically minoritised backgrounds reported racism and cultural and/or religious microaggressions. White British students also experienced discrimination and inequity due to their age, sex, gender, and sexual orientation while also being valued for their whiteness. These experiences impacted whether students felt safe enough to report adverse experiences and whether they felt able to progress in the NHS.

All nursing students were aware of ward culture as a negative workplace environment in which they found themselves. However, feeling unvalued was commonly disclosed by both White British and ethnically minoritised students, but for different reasons. Though White British students were sometimes given tasks that they felt were beneath them, ethnically minoritised students believed that their mentor did not have time to support or guide them and that they were being actively subordinated and therefore, unvalued. These differences in perceptions of value may affect job retention for ethnically minoritised nurses, and whether they consider applying for more senior roles in future. In short, ethnically minoritised student nurses need to feel valued within the workplace for them to remain and progress within the NHS. Limited representation in senior positions may act as a deterrent for potential candidates from an ethnic minority background and prevent nurses from applying for those positions (37). Student nurses who do not encounter professionals from their own racial and ethnic group in leadership positions may feel that there are barriers blocking their own advancement in their careers as less progression is made (38).

This research emphasises the importance of creating supportive and psychologically safe working environments for ethnically minoritised student nurses starting their careers within the NHS. The themes identified in this study support the existence of a hidden curriculum in nursing (27) as an early onset mechanism underpinning the wider culture, one that excludes ethnically minoritised staff and puts them at a disadvantage. This has been found in other recent studies where ethnically minoritised nurses report feeling excluded from opportunities for career progression and *“White networks of power”* (39) and Black African nurses specifically face discrimination, lack of opportunity and poor treatment from staff (19). Other studies have also reported that ethnically minoritised students are aware of the hidden curriculum as a barrier and are inclined to make friendships based on access to the information held in the hidden curriculum to counteract the exclusion (40).

### 5.1. Implications

To disrupt the ingrained hidden curriculum, the implementation of longstanding interventions that reflect and benefit ethnically minoritised students are essential (41). The Hidden Curriculum Evaluation Scale in Nursing Education (HCES-N) should be widely used to determine and evaluate the hidden curriculum in nursing placements (42). In addition, potential reform to the student nurse experience could be achieved by introducing anonymised feedback after each clinical placement, as this would create an ongoing review and capture the learning opportunities provided to all students.

None of the student nurses in this study who reported discrimination had their complaints taken seriously or followed up. This points to a lack of resources and investment in processes of accountability in the NHS more widely. To improve accountability, transparency in reporting incidents of discrimination and the implementation of improved mechanisms for addressing these incidents is crucial. The appointment of a designated person responsible for addressing and resolving incidents of discrimination while establishing a robust tracking system for incidents and monitoring the effectiveness of interventions can further support accountability efforts within the NHS.

In 2019 the NHS began replacing student mentors with practice supervisors, practice assessors, and academic assessors, and all student nurse training programmes required this change to be implemented by September 2020 (43). As these roles are new to the NHS, this is a pivotal time to evaluate their impact and establish effective processes for leadership accountability. These supervisors/assessors should be specifically trained to identify and handle reports of racial discrimination, bullying and harassment and shift the focus away from generic cultural awareness and equality and diversity training, which has been found to be ineffective in tackling discrimination (38, 39). Alternative methods to traditional equality and diversity training, including interactive or experiential training (46) and inclusive leadership training (47), which have been found to be more effective in addressing discrimination in the workplace. Interactive or experiential training involves actively engaging participants in exercises and simulations that simulate real-life situations, allowing them to practice addressing discrimination and other forms of biased behaviour in a controlled environment. Inclusive leadership training teaches leaders and managers how to create a culture of inclusion within their organisations through training on communication, problem-solving, conflict resolution, and the implementation of inclusive leadership practices through coaching and mentorship. Incorporating diversity and inclusion considerations into other types of training and professional development activities, such as leadership development programs, onboarding processes, and performance review systems may also be beneficial.

### 5.2. Limitations

A constraint of this research was that participants were not provided with their transcripts to review or provide feedback, which could have impacted the confirmability of this study. It is important to note that the sample for this study was restricted to London trusts. While it is possible that NHS trusts with lower levels of ethnic diversity may present unique challenges for ethnically minoritised student nurses, such as feelings of isolation, further research is necessary to fully understand the extent of these challenges. Therefore, it is imperative that we also examine the experiences of ethnically minoritised student nurses in trusts outside of London to create initiatives that are appropriate and widely applicable for addressing discrimination within the NHS. As previously demonstrated by Woodhead et al.(8), feelings of isolation may also be a concern for those working in trusts with high levels of diversity but low levels of inclusion, particularly in London.

Further analyses from post-COVID data may offer insight into how students’ experiences of training and career progression was impacted by the COVID-19 pandemic, if/how ward culture changed, and how they navigated burnout, redeployment and psychosocial stress. Nonetheless, findings from this study suggest that these inequalities are present when staff first begin working and training in the NHS and should be used as a starting point to address racial and ethnic inequalities reported by NHS trainees and staff (48).

## 6. Conclusion

These initial experiences of inequality and discrimination are liable to shape a student’s perspective of their profession and ability to progress within nursing. The NHS is responsible for ensuring that student nurses’ developmental opportunities are equal, irrespective of ethnicity. NHS trusts across the UK should focus on the erasure of the barriers faced by racial and ethnically minoritised students and aim to improve the unequal feelings of value during clinical placement. Addressing these tractable problems could help alleviate current retention issues within the NHS.

## Data Availability

All data produced in the present study are available upon reasonable request to the authors

